# The mental health of staff working on Intensive Care Units over the COVID-19 winter surge of 2020 in England: a cross sectional survey

**DOI:** 10.1101/2022.01.13.22269151

**Authors:** Charlotte Hall, Joanna Milward, Cristina Spoiala, Jaskiran Kaur Bhogal, Dale Weston, Henry W. W. Potts, Tristan Caulfield, Michael Toolan, Kate Kanga, Sarah El-Sheikha, Kevin Fong, Neil Greenberg

## Abstract

**Background:** The COVID-19 pandemic generated a surge of critically ill patients greater than the NHS’ capacity. Additionally, there have been multiple well-documented impacts associated with the national COVID-19 pandemic surge on ICU workers, including an increased prevalence of mental health disorders on a scale potentially sufficient to impair high-quality care delivery.

**Aim:** To identify prevalence of probable mental health disorders and functional impairment. As well as establish demographic and professional predictors of probable mental health disorders and functional impairment in ICU staff between November 2020 to April 2021.

**Methods:** English ICU staff were surveyed before, during and after the winter 2020/2021 surge using a survey which comprised of validated measures of mental health.

**Results:** 6080 surveys were completed, by nurses (57.5%), doctors (27.9%), and other healthcare staff (14.5%). Reporting probable mental health disorders increased from 51% (prior to), to 64% (during) and then dropped to 46% (after). Younger, less experienced and nursing staff were most likely to report probable mental health disorders. Additionally, during and after the winter, over 50% of participants met threshold criteria for functional impairment. Staff who reported probable post-traumatic stress disorder, anxiety or depression were more likely to meet threshold criteria for functional impairment.

**Conclusions:** The winter of 2020/2021 was associated with an increase in poor mental health outcomes and functional impairment during a period of peak caseload. These effects are likely to impact on patient care outcomes and the longer-term resilience of the healthcare workforce.

## Introduction

Psychological distress has increased in the general population over the course of the COVID- 19 pandemic^1^ with key workers reporting higher rates of probable mental health disorders than the general population.^2^ Healthcare workers, particularly those working on the frontline, have experienced high rates of mental health challenges such as depression, anxiety, stress, and burnout^3-7^. Furthermore, health and social care workers were already reporting high levels of pre-existing mental health disorders that may have increased their risk of experiencing mental health during a public health emergency.^4^

During the pandemic, staff working on intensive care units (ICUs), including doctors, nurses, and other healthcare professionals, have arguably been the most directly impacted by the surge in critically ill COVID-19 patients. Nurses appear to have been particularly exposed and have reported higher rates of symptoms consistent with common mental disorders and post-traumatic stress disorder (PTSD) compared to other ICU staff.^8^ During the pandemic, ICU staff have faced a constellation of specific stressors. These include the perceived risk to their own health from exposure to COVID-19, very high mortality rates among the patients in their care,^9^ reduced staffing ratios, shortages of personal protective equipment and the need to work beyond their level of seniority.^10^

Poor mental health of ICU staff has the potential to impact the quality and safety of patient care. The phenomenon of presenteeism, in which staff continue to work while functionally impaired by the state of their mental health, may lead to an increased risk of errors and poorer performance, which in turn may impact the quality and safety of patient care.^11, 12^

With COVID-19, and the backlog of care resulting from the pandemic, exerting ongoing pressures on ICU resources, it is important to understand how the mental health of ICU workers has been impacted. This is essential in the identification of risk factors in this population, to help ensure that appropriate support is made available for all,^13^ and to inform future pandemic planning.

Building on the initial ICU mental health survey conducted by Greenberg et al.,^8^ which found substantial rates of probable mental health disorders in ICU staff, this study analysed data from three subsequent timepoints of the survey corresponding to before, during and after the peak of the COVID-19 winter 2020/2021 surge in England, to explore the impact of this surge on the mental wellbeing of staff working in ICUs.^14^

Therefore, the current study aimed to: describe the prevalence of five mental health outcomes: probable depression, probable PTSD, probable general anxiety disorder, and problem drinking, in ICU staff over the winter 2020/2021 surge in England; explore demographic and professional predictors of poorer mental health outcomes in ICU staff over the 2020/2021 winter surge in England; describe the prevalence of functional impairment in ICU staff over the 2020/2021 winter surge in England; and, explore demographic and professional predictors of functional impairment in ICU staff over the 2020/2021 winter surge in England.

## Method

### Study setting

An online cross-sectional survey was designed and run in 56 English ICUs, which experienced a surge in adult patients, above their formally commissioned baseline. Collection occurred across three time points: before the peak - 19^th^ November to 17^th^ December 2020; during the peak - 26^th^ January – 17^th^ February 2021; and after the peak - 14^th^ April – 24^th^ May 2021. These data collection points were part of an ongoing service evaluation of ICU staff’s mental health which commenced in June 2020.^8^

This study was approved by the Psychiatry, Nursing and Midwifery Research Ethics Subcommittee, King’s College London reference number: MOD-20/21-18162.

The 56 NHS hospitals which provided data comprised of District General Hospitals, Teaching Hospitals and Quaternary Paediatric Hospitals. The selection process reflected hospitals utilising surge capacity and hospitals receiving or making use of interhospital transfers as part of mutual aid support between neighbouring units. Where possible, data for hospital baseline ICU bed number (as declared in 2020, immediately prior to the pandemic) and actual maximum occupancy during COVID-19 was collected. All surveyed units exceeded 100% of their baseline ICU capacity during the winter 2020/2021 surge.

### Survey design

Data were collected via an anonymised web-based survey, designed to be completed in less than 5 minutes, comprising validated questionnaires assessing mental health status and psychological well-being. Participants were aware that their participation was voluntary, their data would be anonymised, they were free to stop at any point during the completion of the study and any incomplete surveys would be discarded. The Lime Survey tool (https://www.limesurvey.org/) was used to build the survey and hosted on a dedicated secure university server.

### Survey distribution

Circulation and completion of the survey was encouraged through engagement with clinical leads in each of the intensive care units. The survey was distributed through departmental email and messaging groups. All staff working in ICUs (doctors, nurses, and other healthcare professionals) were eligible to take part. Due to the recruitment method, the size of the sample was determined by the participants who chose to complete the survey. Individual respondents could not be followed across timepoints as the survey was anonymous in order to reduce barriers to reporting.^15,16^ No participant data were excluded. Figure 1 displays a participant flow chart.

**Figure 1.**
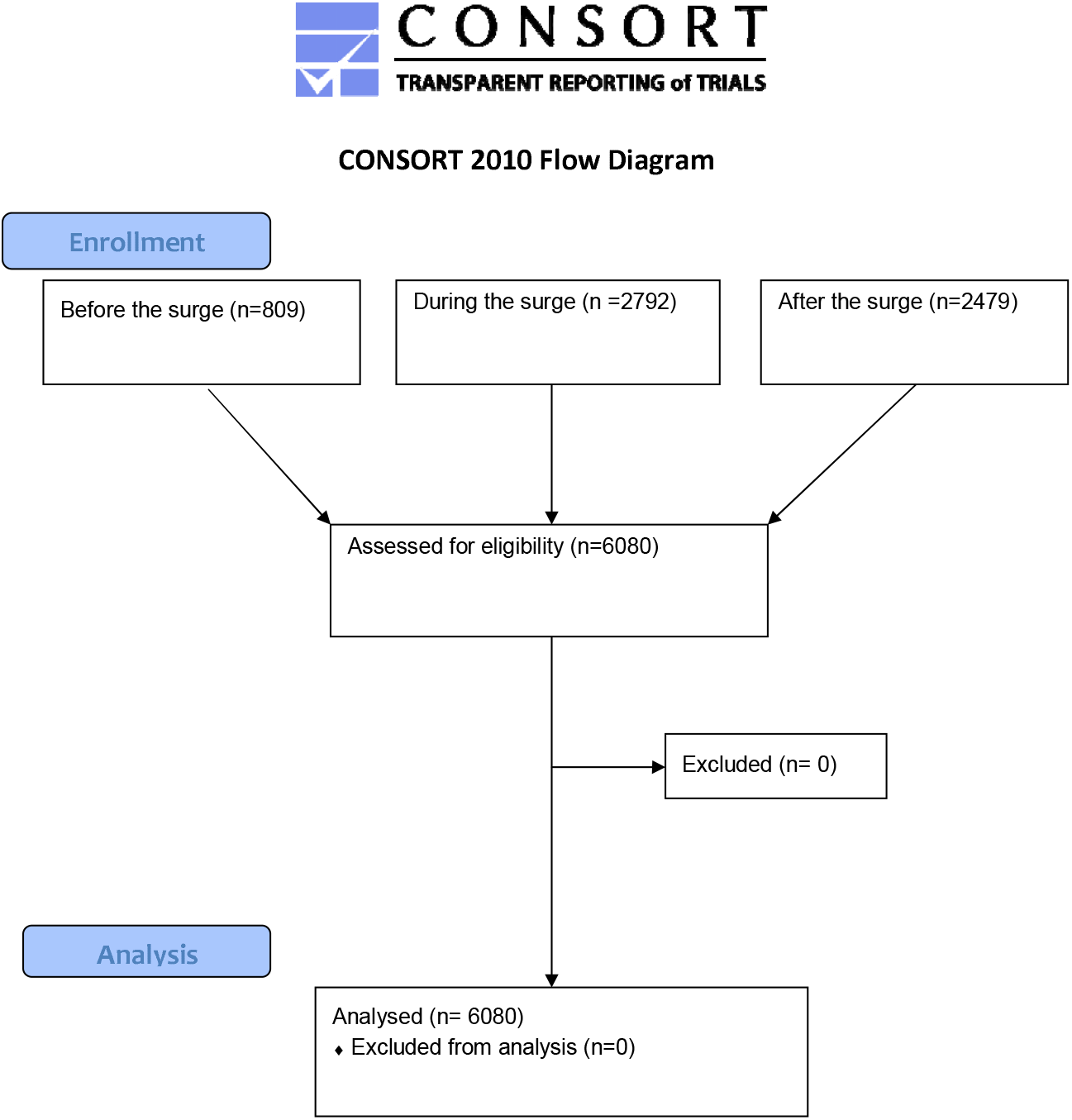
CONSORT 2020 Participant Flow Diagram

### Collected variables and outcome definitions

Demographic data collected included age, gender, job role and seniority. Doctors who were graded FY 1-2, ST 3-4, ST 5-6, ST 6-7 were classed as junior staff (staff still in training) and consultant and senior associate specialists as senior staff. Nurses in Band 5 (i.e. those newly qualified or staff nurses) or Band 6 (i.e. those who are nursing specialists or senior nurses) were classed as junior, with Band 7 (i.e. those who are advanced nurses or nurse practitioners) or higher (e.g. Matrons) classed as seniors.

The following measures, for which binary outcomes were set following cut-off scores to indicate a case, were used; the 9-item Patient Health Questionnaire (PHQ-9) with a score of >9 indicating probable moderate depression and >19 probable severe depression;^17^ the 6-item Post-Traumatic Stress Disorder checklist (PCL-6) with a score of >17 indicating the presence of probable PTSD;^18^ AUDIT-C with a score of >7 indicating problem drinking;^19^ the 7-item Generalized Anxiety Disorder (GAD) scale with a score >9 indicating a probable moderate anxiety disorder and >15 indicating probable severe anxiety disorder.^20^ The primary variable was defined, any mental disorder (AMD), which referred to those meeting the threshold criteria for at least one of the following probable mental disorders: moderate or severe anxiety, moderate or severe depression, problem drinking, or PTSD.

The Work and Social Adjustment Scale (WSAS) was added to the survey during the surge, therefore data is only available for the timepoints during and after the peak. The scale is based on how much an individual’s ability to carry out day-to-day tasks is impacted by an identified problem in their lives (e.g. “*Because of the way I feel my ability to work is impaired*”), and consists of 5 items answered on an 8-point Likert scale. A score of >20 indicated severe psychopathology-related functional impairment and a score of >10 indicated moderate functional impairment.^14^

### Statistics

Using SPSS V27, descriptive statistics were plotted using counts and percentages for all mental health outcomes across the entire sample. The various measures of psychological distress were highly correlated, so one multivariable logistic regression was carried out using AMD, with demographic (i.e. gender, age) and professional variables (i.e. role, seniority) as predictors. A second multivariable logistic regression was carried out for Work and Social Adjustment Scale, with all probable mental health disorders, demographic and professional variables entered as predictors. Because of the small sample size of other healthcare professionals, only doctors and nurses were included in the logistic regressions. Comparator groups were chosen based on expected impact (e.g. junior staff would be impacted more senior staff, so senior staff became the reference category). Additionally, senior nurses were compared to all others (junior nurses and all doctors), and senior doctors were compared to all others (junior doctors and all nurses), as we expected that the effect of seniority might be different across the professions. AMD and WSAS were visually compared across timepoints using forest plots with odds ratios and confidence intervals shown. Inferential statistics comparing across waves were not possible due to lack of independence of observations: as the survey was completed anonymously, we could not match responses in different waves that may have been from the same individuals.

## Results

### Demographics

Table 1 displays the characteristics of the sample used within the current study. Across all three timepoints, most respondents were female, and the modal age group was 30-44 years old. Nurses comprised over 50% of the sample at all timepoints; they were mainly junior (Band 6 or below) and were regular ICU, rather than redeployed, staff. Doctors constituted around 30% of the sample; the majority were anaesthetists and of a senior level (i.e. Senior Associate Specialist or Consultant).

**Table 1.** ICU Participant characteristics.

| Variables | Before the surge<br>(*n* = 809)<br>*n* (%) | During the surge<br>(*n* = 2792)<br>*n* (%) | After the surge<br>(*n* = 2479)<br>*n* (%) |
| --- | --- | --- | --- |
| **Gender** |  |  |  |
| Male | 266 (32.9) | 719 (25.8) | 667 (26.9) |
| Female | 536 (66.3) | 2053 (73.5) | 1790 (72.2) |
| Other<sup>a</sup> | 7 (0.9) | 20 (0.7) | 22 (0.9) |
| **Age** |  |  |  |
| 16-29 | 141 (17.4) | 550 (19.7) | 426 (17.2) |
| 30-44 | 374 (46.2) | 1320 (47.3) | 1216 (49.1) |
| 45-56 | 268 (33.1) | 849 (30.4) | 756 (30.5) |
| 60+ | 26 (3.2) | 73 (2.6) | 81 (3.3) |
| **Role** |  |  |  |
| Doctor | 258 (31.9) | 791 (28.3) | 649 (26.2) |
| Type |  |  |  |
| *Anaesthesia* | 157 (60.9) | 401 (50.7) | 322 (49.6) |
| *ICU* | 89 (34.5) | 317 (40.1) | 280 (43.1) |
| *Other* | 12 (4.7) | 73 (9.2) | 47 (7.2) |
| Grade |  |  |  |
| *Junior*<sup>b</sup> | 93 (36.0) | 300 (37.9) | 197 (30.4) |
| *Senior*<sup>c</sup> | 165 (64.0) | 491 (62.1) | 452 (69.6) |
| Nurse | 428 (52.9) | 1615 (57.8) | 1455 (58.7) |
| Type |  |  |  |
| *ICU* | 351 (82) | 1334 (82.6) | 1260 (86.6) |
| *Other* | 16 (3.7) | 171 (10.6) | 115 (7.9) |
| *Theatres* | 61 (14.3) | 110 (6.8) | 80 (5.5) |
| Grade |  |  |  |
| *Junior*<sup>d</sup> | 329 (76.9) | 1264 (78.3) | 1113 (76.5) |
| *Senior*<sup>e</sup> | 99 (23.1) | 351 (21.7) | 342 (23.5) |
| Other<sup>f</sup> | 123 (15.2) | 386 (13.8) | 375 (15.1) |

**Note:** <sup>a</sup>: indicates both, those who chose to not disclose their gender, and those who selected 'other'. <sup>b</sup>: refers to those who chose the following grading categories: FY 1-2, ST 3-4, ST 5-6, ST 6-7. <sup>c</sup>: refers to those who chose the following grading categories: consultant or senior associate specialist. <sup>d</sup>: refers to those who chose the following grading categories: Band 5 or Band 6. <sup>e</sup>: refers to those who chose the following grading categories Band 7, Band 8 or Band 9. <sup>f</sup>: encompasses the following job roles: Healthcare assistant, Occupational therapist, Operating Department Practitioner (ODP), Pharmacist, Physiotherapist and 'Other'.

### Mental Health Measures

#### Prevalence

Figure 2 shows the percentage of ICU staff meeting the threshold criteria for all tested mental health measures. A clear pattern was observed across the timepoints. The prevalence of all tested mental disorders increased between before and during the peak (e.g. AMD 51.3% [47.8-54.8] *vs* 64.6 [62.8-66.4]), and then decreased after the peak (e.g. AMD 45.5 [43.6- 47.5].

**Figure 2.**
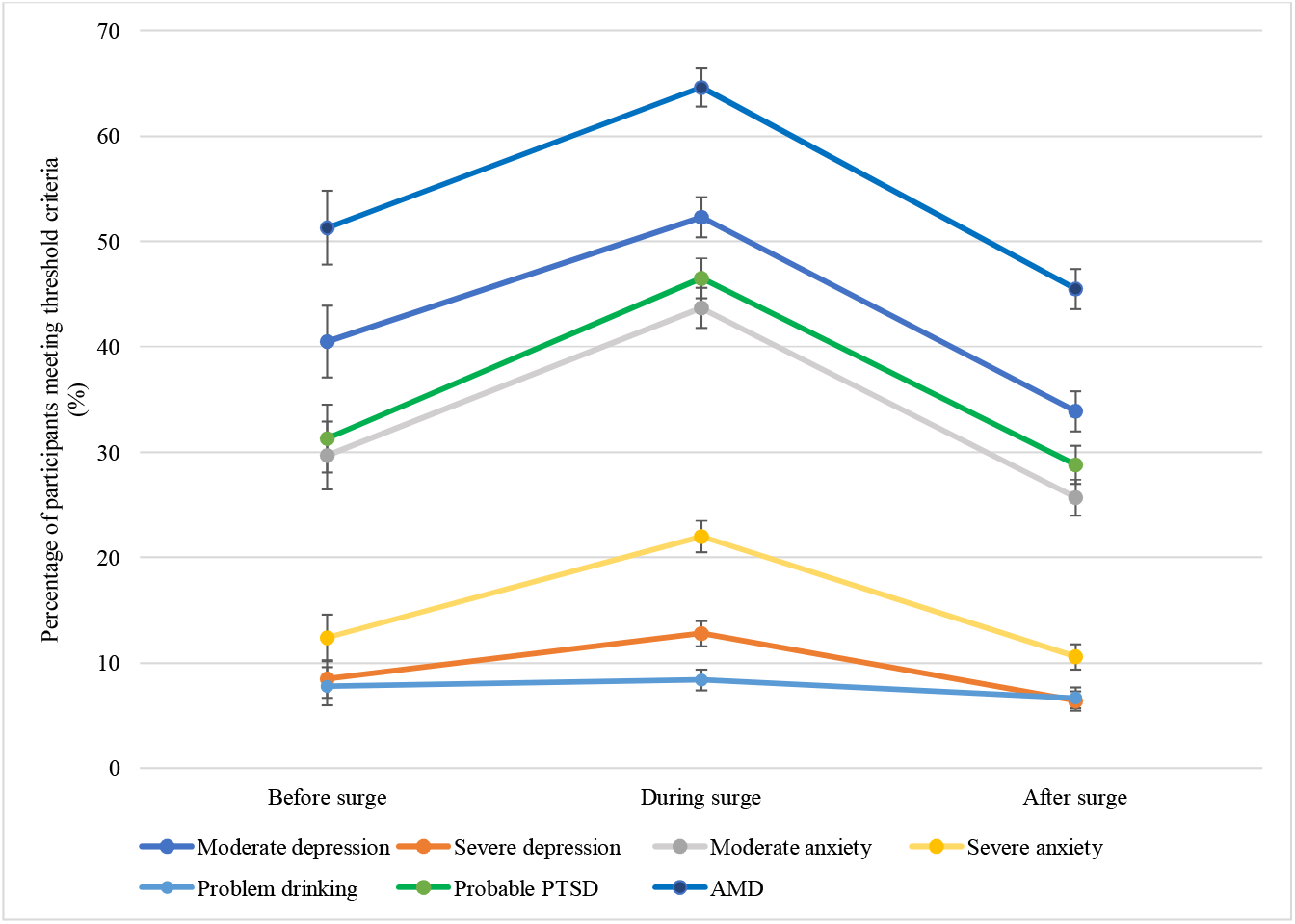
Percentage prevalence and confidence intervals of participants meeting the threshold criteria for depression, anxiety, PTSD and problem drinking across the COVID-19 2020/2021 winter surge. *Note*. Before, after and during samples are independent. The joining lines act as a visual aid. Before surge represents 19th November to 17th December 2020; during the surge represents - 26th January – 17th February 2021; and after the surge represents - 14th April – 24th May 2021.

Probable moderate depression was the most common across all time points (before: 40.5% [37.1-44.0]; during 52.3% [50.4-54.2]; after: 33.9% [32.0-35.8]), followed by probable PTSD (before: 31.3% [28.1-34.6]; during 46.5% [44.6-48.4]; after: 28.8% [27.0-30.6]), and moderate anxiety (before: 29.7% [26.5-33.0]; during 43.7% [41.8-45.5]; after: 25.7% [24.0- 27.5]).

#### Adjusted outcomes

A multivariable logistic regression was performed to ascertain the association of age, gender, job role, and seniority with the likelihood that participants experienced AMD at each of the three timepoints. Results were relatively consistent across time. Figure 3 displays a forest plot of effect size and confidence intervals to allow visual comparison across timepoints. Older staff (30+ years old) showed lower rates of AMD at all timepoints, with this result being statistically significantly for some age groups during and after the peak. Nurses were more likely than doctors to have experienced AMD, although this was only statistically significantly during the peak. Junior nurses were more likely than senior nurses or any doctors to have experienced AMD and this was significant during and after the peak. There were no statistically significant differences by gender or doctor seniority at any timepoint.

**Figure 3.**
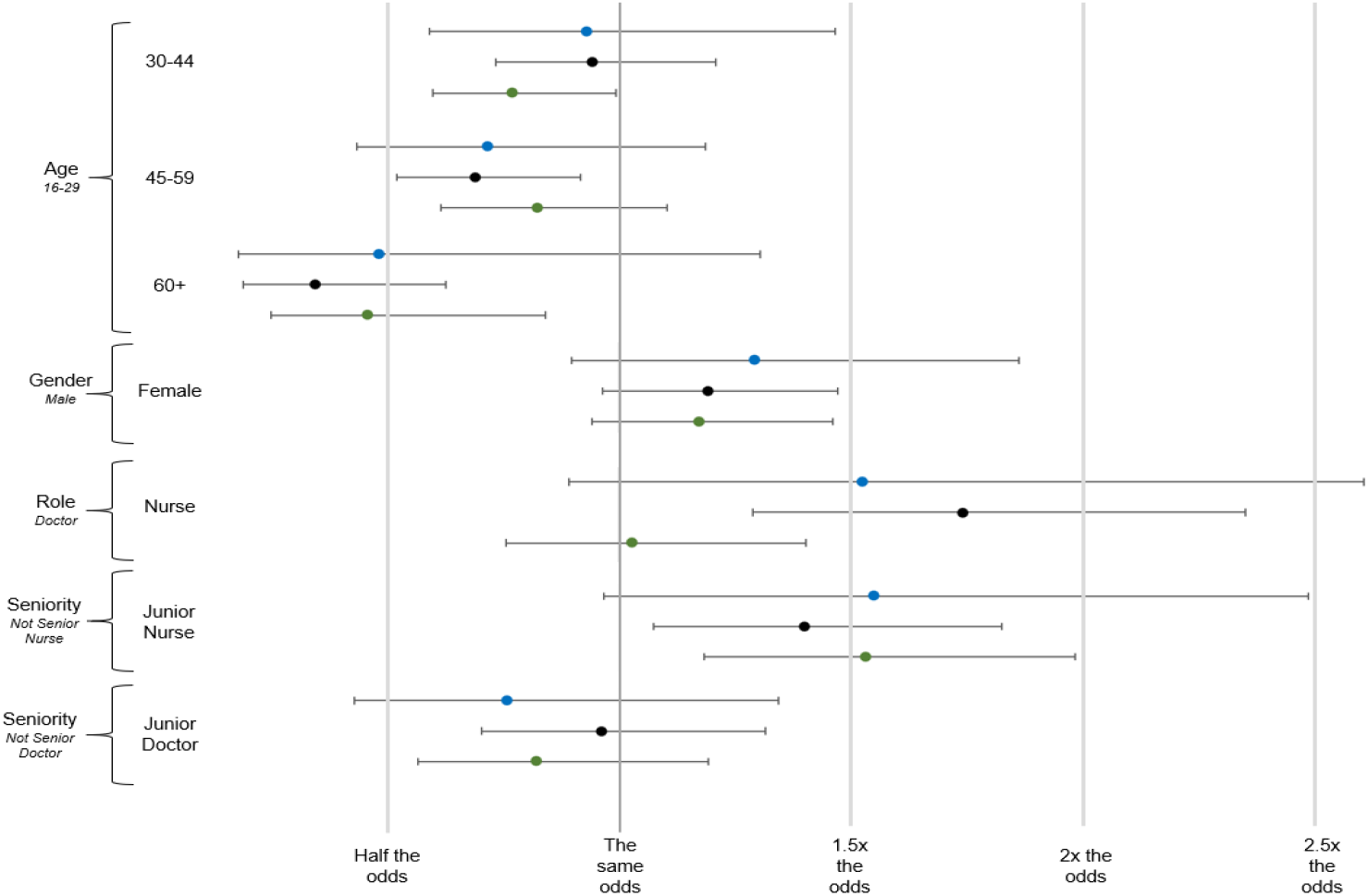
Forest plot displaying confidence internals and effect sizes for each variable’s effect on AMD over each timepoint *Note*: Blue markers indicate before the surge, black markers indicate during the surge, and green markers indicates after the surge. Reference group italicised under each variable. Analysis was only carried out for doctors and nurses, senior nurses were compared to all others (junior nurses and all doctors); senior doctors were compared to all others (junior doctors and all nurses).

### Functional impairment (Work and Social Adjustment Scale)

#### Prevalence

Functional impairment was more prevalent during the peak in comparison to after. During the peak, 69.1% [67.4-70.8] of participants met the threshold criteria for functional impairment (consisting of 27.9% moderate and 41.2% severe). After the peak, 52.8% [50.8-54.7] of participants met the threshold criteria for functional impairment (consisting of 27.3% moderate and 25.5% severe).

#### Adjusted outcomes

A multivariable logistic regression was performed to ascertain the association of age, gender, job role, seniority, and all mental health outcomes, with the likelihood that participants met the threshold criteria for functional impairment at both timepoints. Figure 4 displays a forest plot of effect size and confidence intervals to allow visual comparison across timepoints. Across both timepoints (during and after the peak), those with probable moderate depression (during OR = 4.7, after OR = 4.7), probable moderate anxiety (during OR = 2.4, after OR = 3.3), and probable PTSD (during OR = 6.4, after OR = 4.6) were all more likely to experience functional impairment in comparison to those without. There was no statistically significant relationship with problem drinking. While functional impairment was more prevalent overall during the peak, there was little difference in the likelihood of functional impairment between those with and without AMD (OR = 0.95). After the peak, those respondents with AMD were twice as likely as those without to experience functional impairment. Controlling for mental health outcomes, there were no independent, statistically significant differences by age, gender, job role, or job seniority (for both doctors and nurses) at any timepoint.

**Figure 4.**
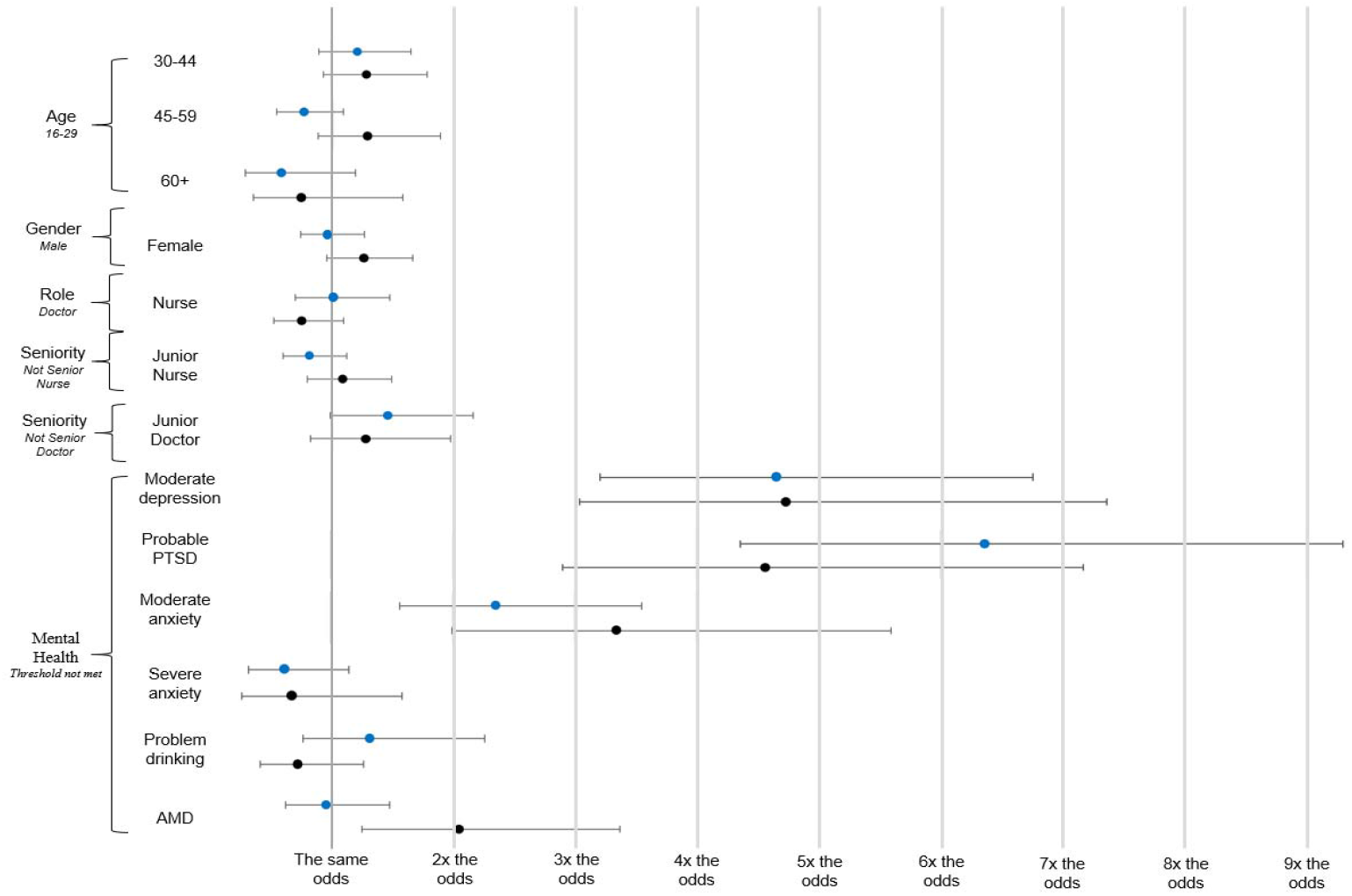
Forest plot displaying confidence intervals and effect sizes for each variable’s effect on functional impairment over each timepoint. *Note*: Reference group italicised under each variable. Analysis was only carried out for doctors and nurses, senior nurses were compared to all others (junior nurses and all doctors); senior doctors were compared to all others (junior doctors and all nurses).

## Discussion

This study examined the mental health of ICU workers between November 2020 to April 2021, during the winter COVID-19 surge in England. At the peak of the winter COVID-19 patient surge, almost two thirds of ICU staff sampled met the threshold criteria for at least one of the surveyed probable mental health disorders. The likelihood of reporting AMD was particularly high in younger, junior nurses. Over half of sampled ICU staff during and after the winter COVID-19 surge met the threshold criteria for functional impairment, with the likelihood of meeting the threshold criteria for functional impairment being substantially increased by the presence of probable PTSD, anxiety or depression.

High rates of probable mental health problems were expected. These findings align with research indicating an increased rate of probable mental health disorders among frontline healthcare staff,^2^ with particular strain during this unprecedented stressful time.^8, 9^ General population studies have shown comparable rates of probable, common mental health problems: using the PHQ-4, Smith and colleagues found comparable case rates and patterns over time in an England population survey,^21^ while Fancourt and colleagues, using the PHQ- 9 and GAD-7, found probable depression and anxiety to be high, but lower over the comparable period.^22^

Beyond the common mental health disorders, our study includes a self-report measure of PTSD symptoms, the PCL-6. We identified that a sizeable fraction of respondents met or exceeded the threshold for probable PTSD at all three time points. Whilst there are no robust pre-pandemic data from ICU staff against which to compare this finding, we note these rates of probable PTSD are comparable to that seen in British military veterans deployed in a combat role during the war in Afghanistan.^23^

Our finding that younger staff were more at risk of reporting AMD was in keeping with previous literature showing similar findings in the general population where younger adults are more likely to report poor wellbeing.^22, 24-27^ However, the risk of reporting AMD was also increased by being a nurse, particularly a junior nurse. This finding matches our previous study, carried out in June/July 2020, which also concluded that nurses were more at risk than other healthcare professionals working in ICU during the COVID-19 pandemic,^8^ as well as other current research.^28,29^ Studies of emergency services^30^ consistently find that lower grade/ranked staff are more likely to report poorer mental health. This may be because those who remain in lower grades are more vulnerable to develop problems in the first place, possibly due to pre-role life adversity which has also been shown to be linked to worse mental health,^31^ or because they are more likely to be directly exposed to significant trauma at work because of their ‘coalface’ role. Similar results may be found for lower grade nursing staff who are more likely to be directly interacting with patients, and relatives, than more senior staff. However, this paper is the first to show a relationship between seniority and mental health among ICU staff.

This study is also the first to examine the relationship between mental health and functional impairment in staff working in ICUs during the COVID-19 pandemic. We found that over half of the participants met the threshold criteria for functional impairment both during and after the peak of the winter 2020/2021 COVID-19 surge. This points to a potential association between poorer staff mental health quality of care and patient outcomes. Indeed a prospective, observational, multicentre study of 31 ICUs reported that depression symptoms were an independent risk factor for medical errors, as were organisational factors such as training and workloads.^32^

Although not causally measured in the current study, the hypothesised associations between functional impairment and patient safety outcomes, which this research points towards, are highly concerning, since safety critical, vigilance tasks are a core feature in the delivery of critical care and thus staff working in ICU settings must function at a high level to ensure the safety and quality of patient care. Mental health status was associated with functional impairment, with those experiencing probable moderate depression, moderate anxiety, or probable PTSD, more likely to meet the threshold criteria for functional impairment, although it is noted that the direction of this relationship was not tested in the current study. The conduct of a study in the context of ongoing, severe COVID-19 patient surge presented myriad challenges. We drew on the experience of other, clinical research teams operating in this environment, and adopted a pragmatic approach to study design, opting for an agile, scalable tool which allowed the capture of data which has clear limitations but nevertheless provides unique insight into mental health impacts on staff during a unique period of operational stress in the NHS. We identified the following principal limitations: Firstly, due to not collecting identifiable data within the surveys (to ensure anonymity), it was not possible to either link cases to allow for longitudinal analysis at the level of individuals, or establish exclusivity between cases, rendering the data collected effectively cross-sectional. Therefore, time (before, after and during the peak) were not entered together into the statistical analysis. Secondly, we do not have data on the current demographic and professional characteristics of the ICU staff population during the COVID-19 crisis, so we do not know how representative the current study is. Additionally, data on ethnicity was not collected as part of the survey, limiting the generalisability of the findings. Thirdly, the recruitment method leaves open the possibility that those with more severe mental health symptoms might be more - or less - likely to participate, thus leading to bias. Fourthly, this study uses self-report measures which only provide an estimate of prevalence; interview-based studies are required to establish the true prevalence of those who would meet diagnostic criteria. Lastly, we recognise that the reported confidence intervals within the regression models are relatively large, which suggests imprecision of observed results. However, this is expected as there were only a limited number of participants at each time point and the differences across time points remain consistent within the confidence intervals, meaning useful conclusions can still be drawn from the analysis.

Future research should explore in further detail the casual relationship between mental health in ICU staff, patient care and outcomes. Such research, into ICU staff’s mental health and functional impairment, should seek to collect identifiable information to allow cases to be linked over time, for a more nuanced statistical analysis to be carried out. Additionally, the Work and Social Adjustment Scale, to measure functional impairment was added to the survey during the surge thus, future additional survey timepoints would allow for further developed analysis of functional impairment.

Recognising that the pandemic placed extraordinary pressure on the NHS, the results of this paper suggest that employers should ensure that all staff working in ICUs are provided with suitable support and this is especially true for more junior nursing staff. While much has been written about how best to support healthcare staff in the workplace (e.g. ^33, 34^), evidence points to promotion of social cohesion at work and its role in reducing PTSD symptoms, such as in a sample of military personnel,^35^ organisational level approaches to help reduce burnout in medics,36 such as changes in schedule and reductions in the intensity of workloads and to ensure that clinical team leaders feel confident to speak to staff about their mental wellbeing.^37^

Whilst the causes of poor mental health and functional impairment in ICU staff during the pandemic are likely to be complex and multifactorial, and determining the causal relationship between them was outside the scope of the current study, it is nevertheless important for healthcare managers to consider strategies to improve the psychological and functional health of their workforce. Delivering high quality care requires functional staff and we suggest that wellbeing initiatives should be seen through the prism of improving patient safety, experience and outcomes and reducing adverse events. In addition to ensuring psychologically healthy workplaces, managers should also take account of the need for strategies such as adequate resourcing and staffing of intensive care units such that individuals reporting high levels of distress can be rested or temporarily rotated away from higher intensity clinical roles. This in turn requires that demand for healthcare services are matched appropriately and realistically with the available supply of staff and resources, although we recognise the exceptional nature of the COVID-19 pandemic made planning and resourcing difficult.

Ultimately, whilst noting caveats about sample representativeness, the current study provides evidence that ICU staff experienced poorer mental health over the winter 2020/2021 COVID- 19 surge with the majority of those surveyed meeting the threshold criteria for poor mental health. Furthermore, this was associated with evidence of high levels of probable functional impairment on a scale that has the potential to negatively impact the safety and quality of patient care. The study also suggests that we should expect staff’s mental health to improve if workload intensity decreases. However, there is, correspondingly, a risk of sustained impairment if demand for healthcare in this setting continues to outstrip capacity. Taken together these findings provide a case for the establishment of a coherent and comprehensive recovery strategy, which appropriately matches demand for healthcare with NHS capacity and human resource, with the goal of protecting staff so that they in turn can continue to deliver safe, high quality patient care. It is essential that staff are properly supported by employers who must recognise the association between mental health status and the ability of staff to safely carry out their caring duties.

## Data Availability

The data used within this study are not publicly available.

## Collaborators

Lindsay Ayres, Emma, Addie, Angharad Williams, Andrew Lynes, Neil Herbert, Clare Chamberlain-Parr, Paula Clements, Peter Hampshire, Maia Graham, Alison Hall, Phoebe Arrowsmith, Rachael Wain, Nadine Weeks, Rosie Holmes, William McCaig, Jessica Miller, Sachin Prabhu, Rebecca Longmate, Sarah Cooper, Paul Stonelake, Maria Crowley, Islam Abdo, Lawrence Wilson, Peter Bamford, Mike Protopapas, Alex Trimmings, Daniel Lutman, Sanjiv Sharma, Dalijit Hothi, Deborah Lees, Anne MacNiven, Bridget Leavey, Ciara McMullin, Dagmar Gohil, Syed Husain, Hozefa Ebrahim, Anil Kambli, Daniel Moult, Maria Mcrittenden, Jane Sansom, Paul Hayden, Susannah Johnson, Akuratiyage Desilva, Nichola White, Sarah Hare, Helen Langrick, Richard Lowsby, Julie Love, Jonathan Phillips, Geradine Hardisty, Jagtar Pooni, Gordon French, Tristan McGeorge, Upeka Ranasinghe, Abdul Nazar, George Collins, Fay Gilder, Rajnish Saha, Sara Blakeley, Catherine Snelson, Meriden Cabales, Deidre McFarlane, Janet Lippett, Paul Dean, Amy Scott, Surrah Leifer, Stephen Krueper, Sandra Barrington, Geeta Aggarwal, Ravi Kumar, Jane Dickson, Edward Cetti, Carole Love, Chris Beevers, Abhijoy Chakladar, Caroline Dean, Dominique Melville, James Nicholson, Aditya Kuravi, Karen Rawlings, Catherine Dexter, Allen Hornby, Andy Higgs, Tim Furniss, Lisa Radley, Laura Langton, Andrew Badacsonyi, Mark Snazelle, Jane Kirk- Smith, Julia Cristall, Karl Woods, Jane Unwin, Anna Dennis, Lisa Millin, Debbie Turner, Nitin Arora, Nick Sherwood, Jonathan Hulme, Rebecca O’Dwyer, Omer Lodi, Ned Hobbs, Manjeet Shehmar, Richard Stewart, Ganesh Suntharalingam, Carol Galvin, Tim Cook, Fiona Kelly, Marghanita Jenkins, Debbie Van Der Velden, Thomas Best, Luis Colorado, Andre Vercueil, Chris Langrish, Elaine Thorpe, Mark Paul, Nick McCartney, Noor Mohammed, James Holding, Shameer Gopal, Jamie Smart, Prabhahar Thaventhran, Richard Breeze, Chris Woolard, Jeremy Wilson, Sinead Hanton, Sean Carroll, Nicholas Barrett, Victoria McCormack, Roopa McCrossan

## Author Contributions

CEH: Performed data analysis, drafted the manuscript, constructed all tables, designed all figures and prepared the manuscript for submission. JM: Study coordinator, developed protocol, supported data analysis, contributed to article revisions. CS: Contributed to protocol, supported data analysis, and contributed to article revisions. JKB: Contributed to protocol, contributed to write-up and article revisions. DW: Provided feedback on protocol and article revisions. HWWP: Supported and guided data analysis, commented on multiple versions of the draft manuscript. TC: Designed the electronic survey tools, supported data analysis and contributed to article revisions. MT: Assisted with recruitment and data collection, contributed to study design and article revisions. KK: Assisted with recruitment and data collection, contributed to study design and article revisions. SES: Assisted with recruitment and data collection, contributed to study design and article revisions. KF: Initiated the concept and formulated the initial design of the study and was a senior advisor to the project. NG: Led study design, study deployment and study team, contributed to serial article revisions. All authors have commented earlier versions of the manuscript and read and approved the final version of the manuscript.

## Data Sharing

The data used within this study are not publicly available.

